# Genetic and behavioural architecture of childhood eating behaviour and links to obesity risk

**DOI:** 10.64898/2026.09.02.26362007

**Authors:** R Karimi, M Baur, GM Power, J Sundfjord, N Fragoso-Bargas, L Clement, OA Andreassen, G Davey Smith, PR Njølstad, RE Brandlistuen, H Ask, G Hemani, KK Ong, Z Kutalik, A Havdahl, M Vaudel, S Johansson

## Abstract

**Background/Objectives:** Childhood appetitive traits are heritable behavioural phenotypes hypothesized to link genetic susceptibility to obesity risk. Yet their genetic architecture and role in mediating polygenic adiposity risk remain poorly understood.

**Methods:** We conducted the largest survey of childhood eating behaviour to date, allowing us to perform genome-wide association studies of six appetitive domains derived from 18 items of the parent-reported Children’s Eating Behaviour Questionnaire in up to 31,018 eight-year-old children from the Norwegian Mother, Father and Child Cohort Study (MoBa). A trio-based design enabled decomposition of direct and indirect genetic effects on appetite and BMI.

**Results:** We identified ten independent genome-wide significant loci for childhood eating behaviour, primarily across *Food Responsiveness*, *Satiety Responsiveness*, and *Food Fussiness*, eight of which lie at established childhood or adult BMI loci. *Food Responsiveness* and *Satiety Responsiveness* showed both phenotypic and genetic correlations with BMI trajectories from early childhood through adolescence. Statistical mediation analyses indicated that 22.1% and 10.4% of the aggregated genetic association with BMI at age 8 could be decomposed through these traits, respectively. Locus-specific patterns further suggested mechanistic pathways, with the *FTO* locus acting predominantly via *Food Responsiveness*, and the *ADCY3* locus via *Satiety Responsiveness*. Trio analyses demonstrated that both BMI and eating behaviour associations were predominantly explained by children’s inherited alleles, with minimal contribution from indirect effect from parental adiposity, although parental genetic liability influenced reporting of *Satiety Responsiveness*.

**Conclusions:** Childhood appetitive traits capture a substantial proportion of genetic susceptibility to adiposity through distinct eating behaviour pathways (under standard mediation assumptions). These effects are primarily driven by the child’s own genotype rather than indirect parental influences, positioning appetite as a plausible, biologically grounded target for early obesity prevention.

## Introduction

Childhood obesity is one of the most pressing public health challenges of our time, with genetic susceptibility playing a central role in shaping individual risk. Yet the pathways through which common genetic variants translate into excess weight gain remain incompletely understood. The Behavioural Susceptibility Theory of obesity proposes that individual differences in appetitive traits act as key behavioural intermediaries linking genetic predisposition to weight gain in an obesogenic environment (Carnell & Wardle, 2007^1^; 2008^2^). These traits encompass behavioural tendencies that influence when, why, and how much individuals eat, including heightened *Food Responsiveness*, reduced *Satiety Responsiveness*, and *Slowness in Eating*. Such traits are well-captured by the Child Eating Behaviour Questionnaire (CEBQ), a validated parent-report instrument that reliably measures distinct appetitive domains in childhood (Wardle *et al*., 2001^3^).

Twin and family studies consistently demonstrate that many childhood appetitive traits are highly heritable, with estimates typically ranging from 60–80% across infancy and childhood (Carnell *et al*., 2008^4^; Dubois *et al*., 2013^5^). Longitudinal prediction studies further show that these traits prospectively predict weight gain and BMI trajectories independent of baseline adiposity (van Jaarsveld *et al*., 2011^6^; 2014^7^). Cross-sectional and prospective studies further show that food approach traits, particularly *Food Responsiveness*, are positively associated with BMI across childhood and adolescence, while food avoidant traits such as *Satiety Responsiveness* and *Slowness in Eating* are inversely associated with adiposity (Kininmonth *et al*., 2021^8^). Taken together, this evidence positions eating behaviours as stable, heritable behavioural phenotype that shape long-term growth trajectories, yet the molecular genetic architecture underlying individual differences in these traits has remained almost entirely uncharacterised.

Genome-wide association studies (GWAS) have identified numerous loci contributing to childhood and adult BMI, many of which implicate appetite-regulating hypothalamic and reward circuits (Bradfield *et al*., 2019^9^; Vogelezang *et al*., 2020^10^; Helgeland *et al*., 2022^11^). Variants near *FTO*, for example, are among the most robustly replicated obesity loci and have been functionally linked to altered food cue reactivity and increased energy intake in children (Frayling *et al*., 2007^12^; Wardle *et al*., 2009^13^; Timpson *et al*., 2008^14^). Similar observations have been made for loci near *MC4R* and *ADCY3*, components of the leptin–melanocortin pathway and ciliary-based satiety signalling in the hypothalamus (Stutzmann *et al*., 2009^15^; Siljee *et al*., 2018^16^). Whether these loci act on adiposity through shared or distinct appetitive mechanisms is unknown.

Children inherit obesity-predisposing alleles from their parents, but parental genetic liability may also indirectly shape the child’s eating environment through feeding practices, food availability, and the role modelling of eating behaviour; a phenomenon termed genetic nurture (Kong *et al*., 2018^17^). Disentangling whether parent-reported child appetite reflects the child’s own inherited biology, parental indirect genetic effects, or parental perceptual biases would allow distinguishing the innate vs. acquired risk of obesity.

Here, we investigate six appetitive domains — *Food Responsiveness (FR), Satiety Responsiveness (SR), Food Fussiness (FF), Slowness in Eating (SiE), Emotional Overeating (EO),* and *Emotional Undereating (EU)* — and their individual items (Table S1) captured with the CEBQ in up to 31,018 eight-year-old children from the Norwegian Mother, Father and Child Cohort Study (MoBa) to study childhood eating behaviour at genome-wide scale. We characterise the phenotypic and genetic architecture of these appetitive traits, identify the behavioural pathways through which common BMI loci exert their effects, and determine the extent to which parent-reported appetite reflects the child’s own inherited biology versus parental influence.

## Results

### 1. Psychometric Validation, Distributions, and Phenotypic Correlations of the Short-Form CEBQ

Using a short-form version of the CEBQ available from MoBa, we derived six composite domains of appetitive behaviour from parent-reported item responses in up to 31,018 children at age 8 years (Figure 1). All domains showed generally acceptable to strong internal consistency (α = 0.70–0.92). Parallel analysis supported retention of six factors, and the six-domain structure was reproduced in an independent confirmatory sample, with standardized CFA loadings ranging from 0.56 to 0.98, supporting their use as quantitative phenotypes for downstream genetic analyses (Table S2). Response distributions were domain-specific: FR and EO were skewed toward lower categories (“Never”– “Seldom”), whereas the remaining domains showed broader distributions centred around mid-scale responses (“Seldom”– “Often”) (Figure 1; Table S2).

**Figure 1.**
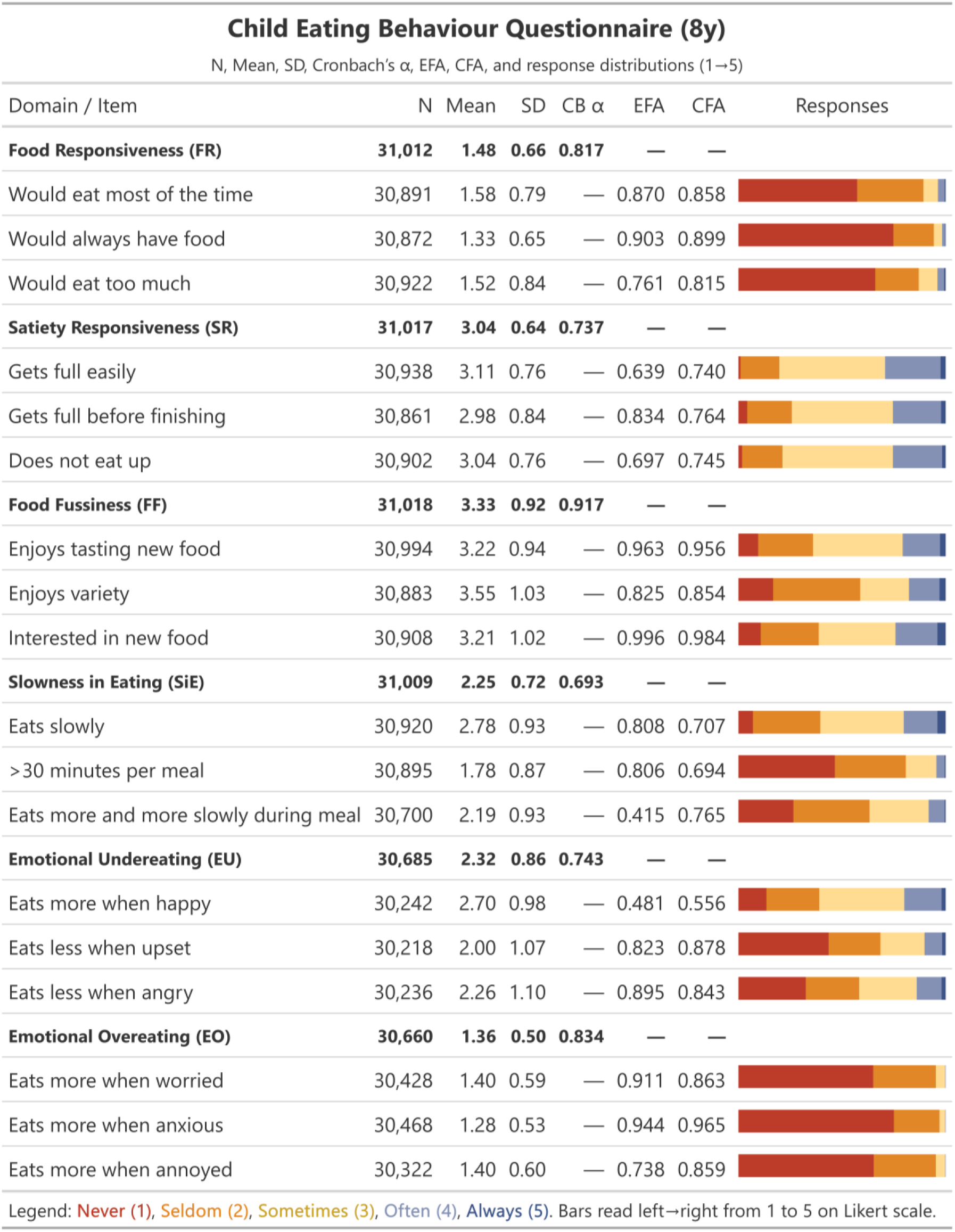
Overview of Childhood Eating Behaviour Questionnaire ( CEBQ) domains and items in the Norwegian Mother, Father and Child Cohort Study (Mo Ba) at age 8 years. Items were rated by parents on a 5 -point Likert scale (1 = Never [red], 2 = Seldom [orange], 3 = Sometimes [yellow], 4 = Often [light blue], 5 = Always [dark blue]). N denotes the number of available responses for the corresponding domain or item. Cronbach’s α reflects internal consistency across the three items within each domain. EFA and CFA values represent standardized factor loadings from the independent exploratory (N = 14,292) and confirmatory (N = 14,288) subsamples, respectively. See Table S2 for full item-level response distributions and factor-model fit statistics.

To characterize developmental and familial associations between child appetite at age eight years and adiposity, we computed Pearson correlations between the six domains, 13 standardized measures of mother-reported BMI from birth to 14 years, and parental pre-pregnancy BMI (Figure 2; Tables S3–S5). *Food Responsiveness* was positively correlated with BMI from birth (r = 0.06), increased across childhood, peaked at ages 7–8 years (r = 0.40– 0.41), and remained evident at age 14 years (r = 0.30). FR was also positively associated with maternal and paternal pre-pregnancy BMI (r = 0.11 and 0.09, respectively). *SR* showed the opposite pattern, with inverse correlations across all ages that were strongest at 7–8 years (r = −0.26). *FF*, *SiE*, and *EO* showed weaker associations with BMI, whereas *EU* was largely null (Figure 2A; Table S3). Between appetitive domains, *FR* correlated positively with *EO* (r = 0.47), while *SR* correlated inversely with *FR* (r = −0.25) and positively with *SiE* (r = 0.36) (Figure 2B; Table S4). Within-domain item correlations were strongest for *FF*, *FR*, and *EO*, and more modest for *SR*, *SiE*, and *EU* (Figure 2C; Table S5).

**Figure 2.**
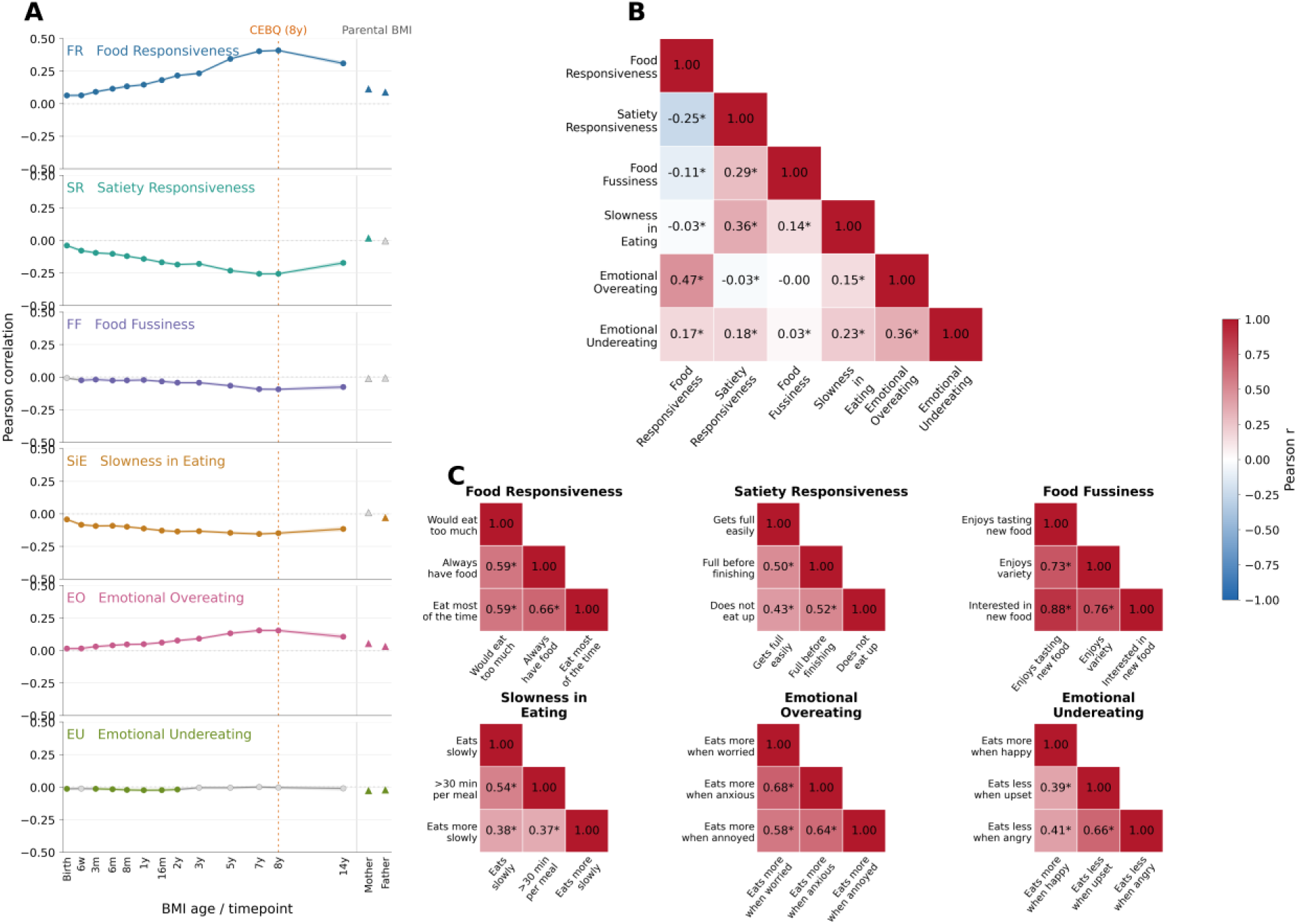
Phenotypic correlations of childhood appetite traits and adiposity in the Mo Ba cohort. **(A)** Pearson correlations between CEBQ domains at age 8 years and standardized BMI from birth to 14 years, including parental pre-pregnancy BMI. **(B)** Pairwise correlations among the six CEBQ domains. **(C)** Item-level correlations within each domain. Colour scale indicates Pearson’s r.

Because appetite and BMI were measured concurrently at age 8 years, we also compared the observed appetite–BMI correlation with a benchmark correlation predicted from two quantities: the correlation between BMI at that age and BMI at age 8, and the correlation between BMI at age 8 and the appetite domain. This benchmark represents the correlation pattern expected if associations at other ages were explained only by BMI tracking through age 8. Longitudinal associations, particularly for *FR*, *SR*, and *SiE*, remained evident beyond this BMI-at-8 benchmark, suggesting that they were not solely driven by concurrent BMI at the appetite assessment (Table S3, Figure S1).

### 2. Single nucleotide polymorphism (SNP) - based Heritability and Genetic Architecture of CEBQ Domains

We performed GWASs for the six domains and their individual items (18 items). SNP-based heritability (h2) estimates were strongest for *Food Fussiness* (h² = 0.18, 95% CI: 0.14–0.22), *Satiety Responsiveness* (h² = 0.15, 95% CI: 0.11–0.19), *Slowness in Eating* (h² = 0.11, 95% CI: 0.07–0.14), and *Food Responsiveness* (h² = 0.10, 95% CI: 0.07–0.13), while heritability estimates for emotional eating traits crossed the null. Item-level heritability was generally comparable to domain-level traits, although *FR-item “If allowed to, my child would eat too much”* and *SR-item “My child gets full easily”* showed higher estimates within their domains (Figure 3A; Table S6).

**Figure 3.**
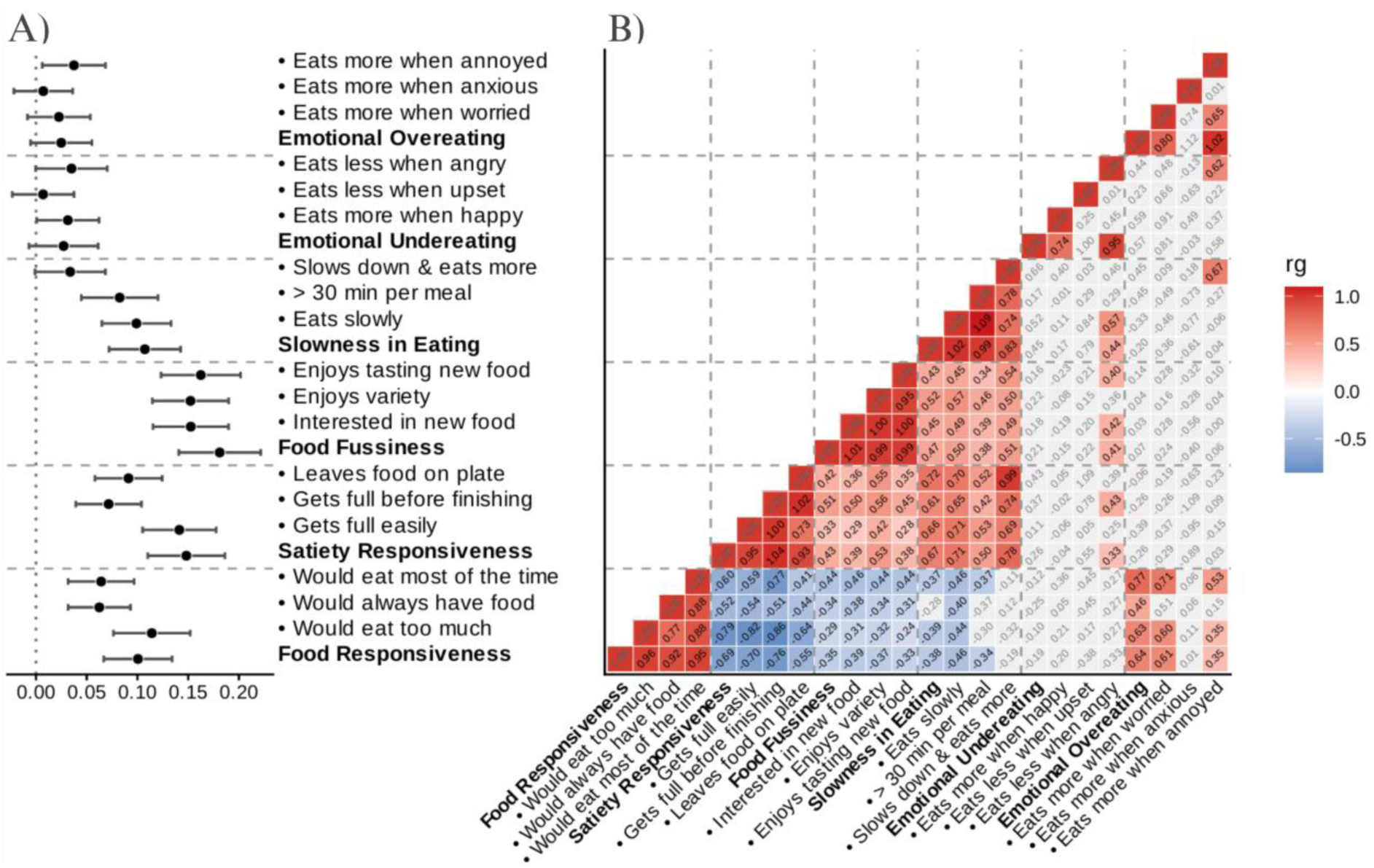
Heritability and genetic correlations within the domains. **A)** SNP-based heritability estimates for each CEBQ domain and individual items (•). **B)** Genetic correlation heatmap, generated from a total of 24 genome-wide association studies (GWAS) for each of the six domains and 18 items (•). MoBa cohort – CEBQ 8 years – SNP-based heritability (h²).

Pairwise genetic correlations broadly mirrored phenotypic patterns, albeit with generally stronger correlations. *FR* was negatively correlated with *SR* (r_g_ = −0.69) and *SiE* ( r_g_ = −0.38), while *FF* was negatively correlated with *FR* (r_g_ = −0.35) and positively correlated with *SR* (r_g_ = 0.43) and *SiE* (r_g_ = 0.47). *SR* and *SiE* were also positively correlated (r_g_ = 0.67). *EO* showed a positive correlation with *FR* (r_g_ = 0.64), whereas *EU* showed no consistent association (Figure 3B; Table S7). Overall, these patterns are consistent with a separation between food-approach (*FR, EO*) and food-avoidant (*SR, SiE, FF*) behaviours. Within-domain genetic correlations were uniformly strong across FR, SR, FF, and SiE (Figure 3B), while emotional-eating composites could not be evaluated because of low heritability.

### 3. Genetic Correlations Between Appetite Traits and BMI Across the Lifecourse

We estimated genetic correlations between the six domains and BMI from birth to age 14 years using LD score regression, including comparative body size at age 10 (SAC10) (Richardson *et al*., 2020^18^) and adult BMI (Yengo *et al*., 2018^19^) (Figure 4, Table S8). Genetic correlations largely reflected the phenotypic correlations reported above albeit with often substantially stronger correlations. *FR* showed the strongest and most consistent positive correlations with BMI, increasing from infancy, peaking around 7–8 years (r_g_ = 0.92–0.98), remaining high at SAC10 and 14 years, and attenuating into adulthood. In contrast, *SR* and *SiE* showed increasingly negative correlations across childhood, strongest in later childhood (e.g. at 8 years, *SR*: r_g_ = −0.73; *SiE*: r_g_ = −0.50). *FF* showed only modest negative correlations with BMI despite its relatively high SNP heritability, whereas *EU* showed no consistent associations. *EO* showed positive correlations that were less precisely estimated, consistent with its lower heritability. *FR-item* “*If allowed to, my child would eat too much”* and *SR-item* “*My child gets full up easily”* showed stronger correlations with BMI than their respective domain composites (Figure S2), indicating that these items capture particularly informative aspects of adiposity-related appetite.

**Figure 4.**
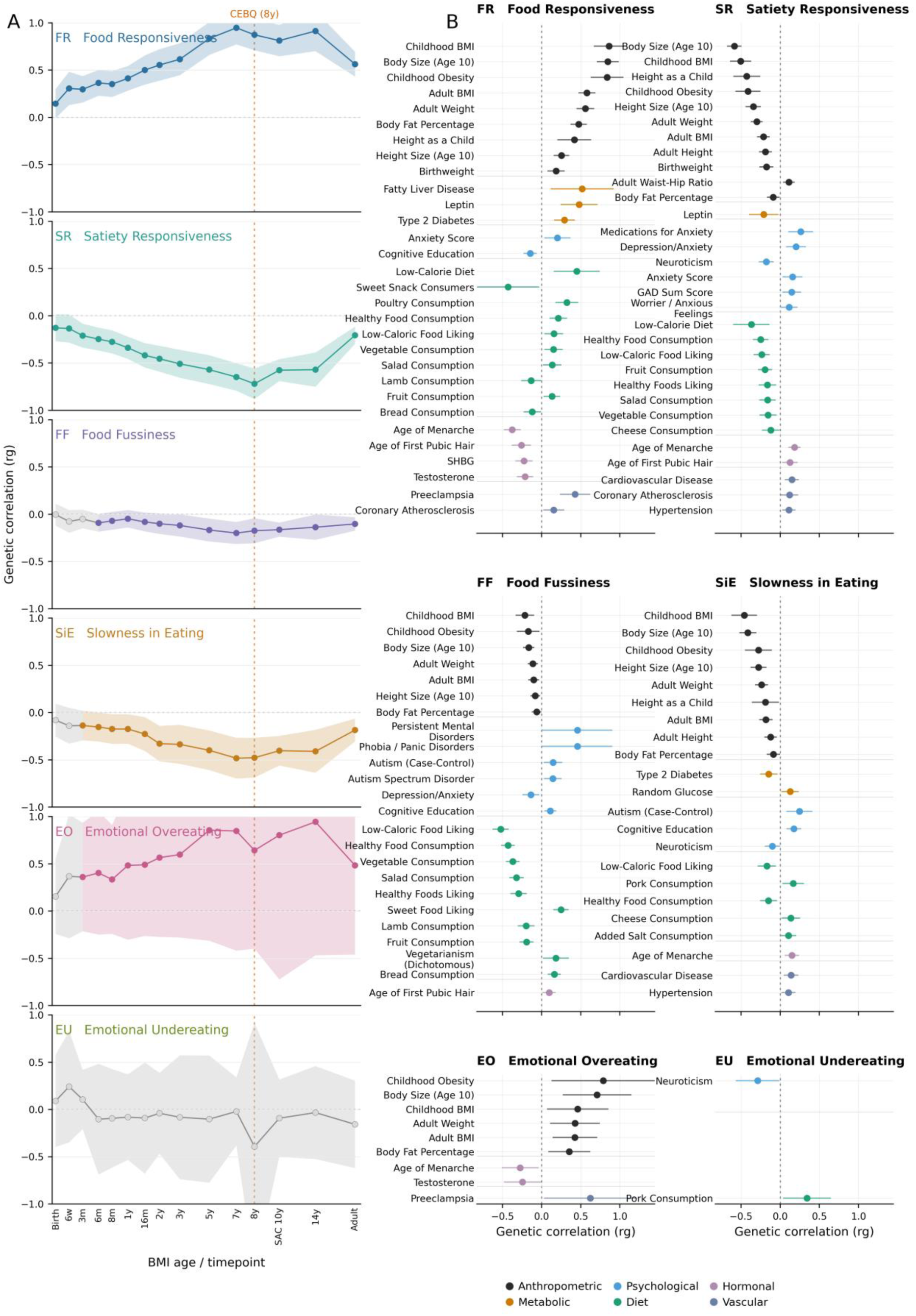
Genetic correlation between A) early-life body mass index ( BMI) signals and the six domains of the Childhood Eating Behaviour Questionnaire ( CEBQ) and B) external GWAS results from anthropometric, metabolic, dietary, hormonal, cardiovascular, and psychological traits. Panel B shows FDR-significant genetic correlations only. Shaded ribbons denote 95% confidence intervals and the dashed line marks age 8 (CEBQ assessment).

To place these findings in a broader biological context, we further estimated genetic correlations with a wide panel of external GWAS results from anthropometric, metabolic, dietary, hormonal, cardiovascular, and psychological traits (Figure 4B, Table S9). *FR* showed strong positive correlations with childhood and adult adiposity, body fat, leptin, and type 2 diabetes, alongside earlier pubertal timing; *EO* showed a similar but less precisely estimated pattern. In contrast, *SR* and *SiE* displayed inverse correlations with adiposity-related traits and later pubertal timing. *FF* was genetically distinct, showing stronger correlations with food preference and aversion traits and comparatively weaker links to adiposity, alongside positive correlations with neurodevelopmental traits.

### 4. Identification of Genetic Loci Associated with Eating Behaviour

We identified ten independent signals at nine genomic loci reaching genome-wide significance (p < 5×10^-8^) across three of the six domains, and their items, within *FR*, *SR*, or *FF* (Figure 5A, Figure S3 & Table S10). Eight are located near regions previously associated with childhood or adult BMI, with several variants showing associations spanning appetite domains and life course BMI profiles (Figure 5B, Table S11). The strongest genetic associations were observed for items in the *Food Responsiveness* domain, with eight loci identified across domain and item-level analyses including variants near *FTO* (p = 6.08×10⁻²³), *SEC16B* (p = 3.47×10⁻²¹), *MC4R* (p = 4.40×10⁻¹¹), *TMEM18* (two independent signals: rs77165542, p = 1.66×10⁻⁹; rs2867131, conditional p = 4.34×10⁻⁸), *TFAP2B* (p = 9.74×10⁻⁹), *KDM4C* (p = 1.10×10⁻⁸), and *ADCY3* (p = 4.77×10⁻⁸). All eight loci were independently significant in the single item “*If allowed to, my child would eat too much”*, while the remaining two *FR* items captured only the *FTO* signal at GWAS significance through colocalizing SNPs (Figure 5A; Table S10).

**Figure 5.**
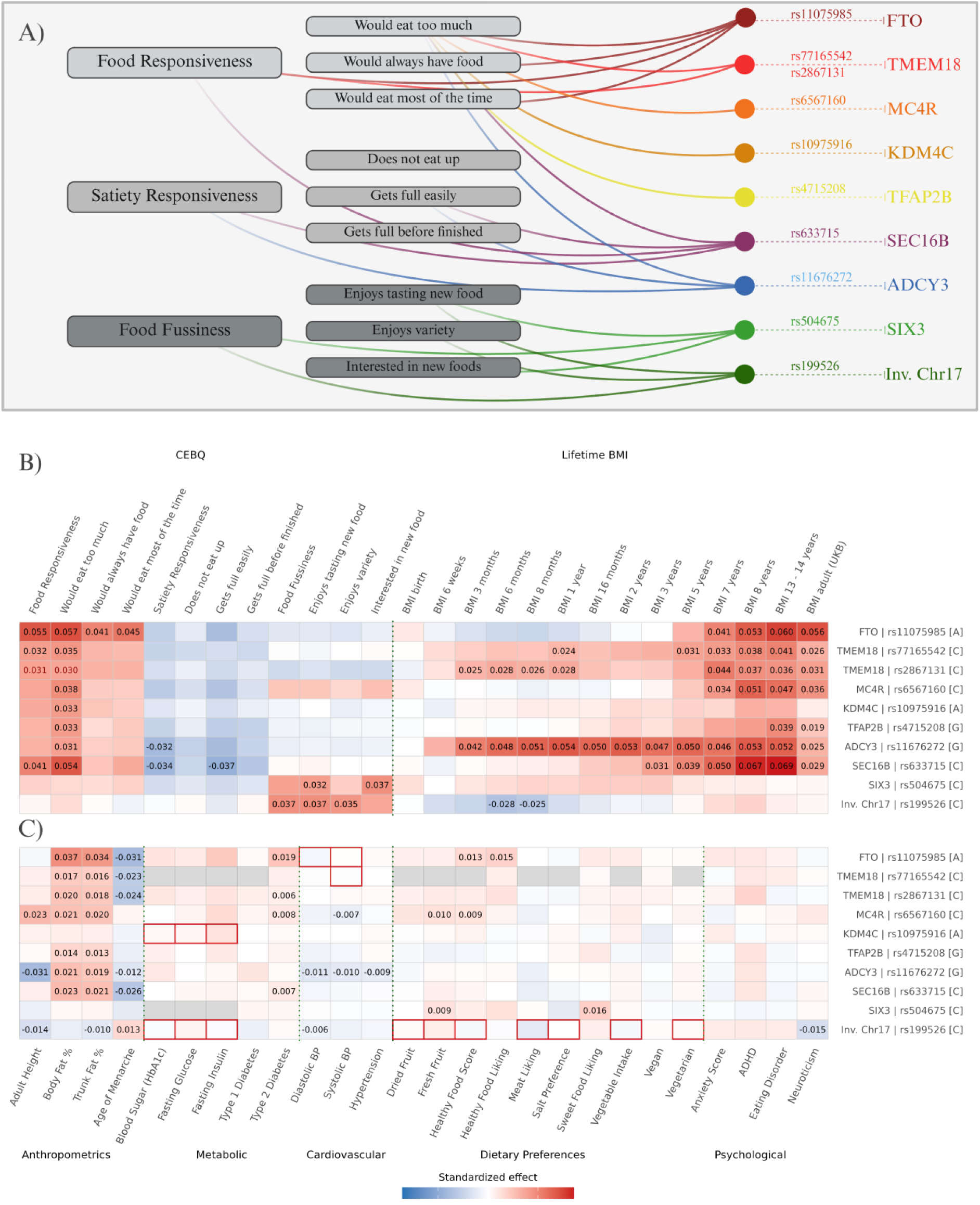
A) Graphical overview of genetic associations for the eating behaviour questionnaire (CEBQ) items and domain scores, including annotations of lead SNPs (after COLOC analysis) and nearest gene annotation and B) effect size heatmaps showing sample-size-adjusted standardized genetic effects, calculated as 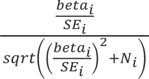, to facilitate comparison across GWAS with differing sample sizes. Effects are shown for all ten genome-wide significant signals identified across 24 GWAS for the individual childhood CEBQ domains and items, as well as early childhood body mass index ( BMI) at 13 consecutive timepoints from birth to 8 years of age + self-reported size at age 10 from UK biobank ^18^ and adult BMI ^19^. C) Selected GWAS Catalog traits for overarching traits of food liking, behavioural and neurological phenotypes, as well as metabolic diseases and diabetes, blood pressure and glucose measurements. Tiles with numerical values indicate studies where this rsID reached genome - wide significance (P<5×10 - 8). Red borders highlight cases in which the lead SNP was not available in the original summary statistics and was substituted with a proxy variant (r² > 0.8). Gray tiles indicate that neither lead nor proxy variants, were available for this study.

For *Satiety Responsiveness*, lead signals near *SEC16B* (p = 1.93×10⁻¹⁰) and *ADCY3* (p = 2.93×10⁻⁸) reached genome-wide significance at both the domain level and for the item “*My child gets full easily”* (Figure 5A; Table S10). Despite that both loci are also associated with *FR*, the two *SR* signals otherwise showed distinctly different multi-trait profiles (Figure 5C, Table S11): The SR-decreasing allele at the *ADCY3* signal was associated with higher BMI already from infancy, shorter adult height, and lower blood pressure, whereas the *SEC16B SR*-decreasing allele began showing associations with higher BMI from age three years and onwards, had stronger effect on earlier menarche, and increased risk of type 2 diabetes. In contrast, the *FTO* locus with comparable strong association with BMI at age 8 years did not reach genome-wide significance for *SR* suggesting that BMI-associated loci do not act uniformly across appetitive traits consistent with *FTO* primarily influencing food approach and *ADCY3* showing stronger association with Satiety-related mechanisms.

*Food Fussiness* showed a genetically distinct architecture, with two significant loci at *SIX3* (p = 1.79×10⁻¹⁰) and the *17q21.31 inversion polymorphism* (rs199526, p = 1.05×10⁻¹⁰). Neither locus has been identified in childhood BMI GWAS at age 8 years, although the inversion locus shows a negative association with early-life BMI (6 months–1 year) in MoBa (Figure 5C) (Helgeland *et al*., 2022^11^). The strength of associations at these loci were broadly consistent across items. *SIX3* showed positive associations with food liking traits, whereas the inversion locus was linked to neurodevelopmental traits, consistent with the broader genetic correlation profile of *Food Fussiness* and supporting a role in food preference rather than adiposity (Figure 5C, Table S11).

Adjustment for BMI at age 8 attenuated the effect estimates for *FR-* and *SR-*associated loci by ∼31–76%, although most signals remained nominally significant after adjustment. The strongest attenuation was observed for the *FR*-*ADCY3* in association (76.3%), which was no longer significant after adjustment (p = 0.191) consistent with substantial overlap between the ADCY3-*FR* association and adiposity at this age, whereas the attenuation for *SR* was less (48%), suggesting a comparatively stronger BMI-independent component. In contrast, *FF*-associated loci showed no attenuation, suggesting relative independence from concurrent adiposity (Table S12; Figure S4).

### 5. Classification of Established BMI Loci

We next sought to systematically classify established childhood BMI loci according to their relative association with *FR* and *SR*. We used 23 loci from the largest childhood BMI GWAS not including MoBa^10^, of which six colocalized with the lead SNPs for *FR* or *SR*. We assessed their association with appetite and BMI in MoBa at age eight together with the two additional *BMI*-loci identified in MoBa at age 8 (*KDM4C* and *TMEM18*) (Figure 6, Table S13). Across SNPs, the estimated effects on *FR* and *SR* correlated positively and negatively, respectively, with effects on BMI (Figure 6A), suggesting that established childhood BMI loci are associated with both food approach and satiety-related appetitive domains. We performed a principal component analysis of effect estimates across BMI, *FR*, and *SR* for all 25 loci (Fig 6B-C, Table S13). The first principal component (PC1), explaining 84% of the variance, reflected overall BMI effect size, whereas the second component (PC2) separated loci according to their relative influence on *FR* versus *SR*.

**Figure 6.**
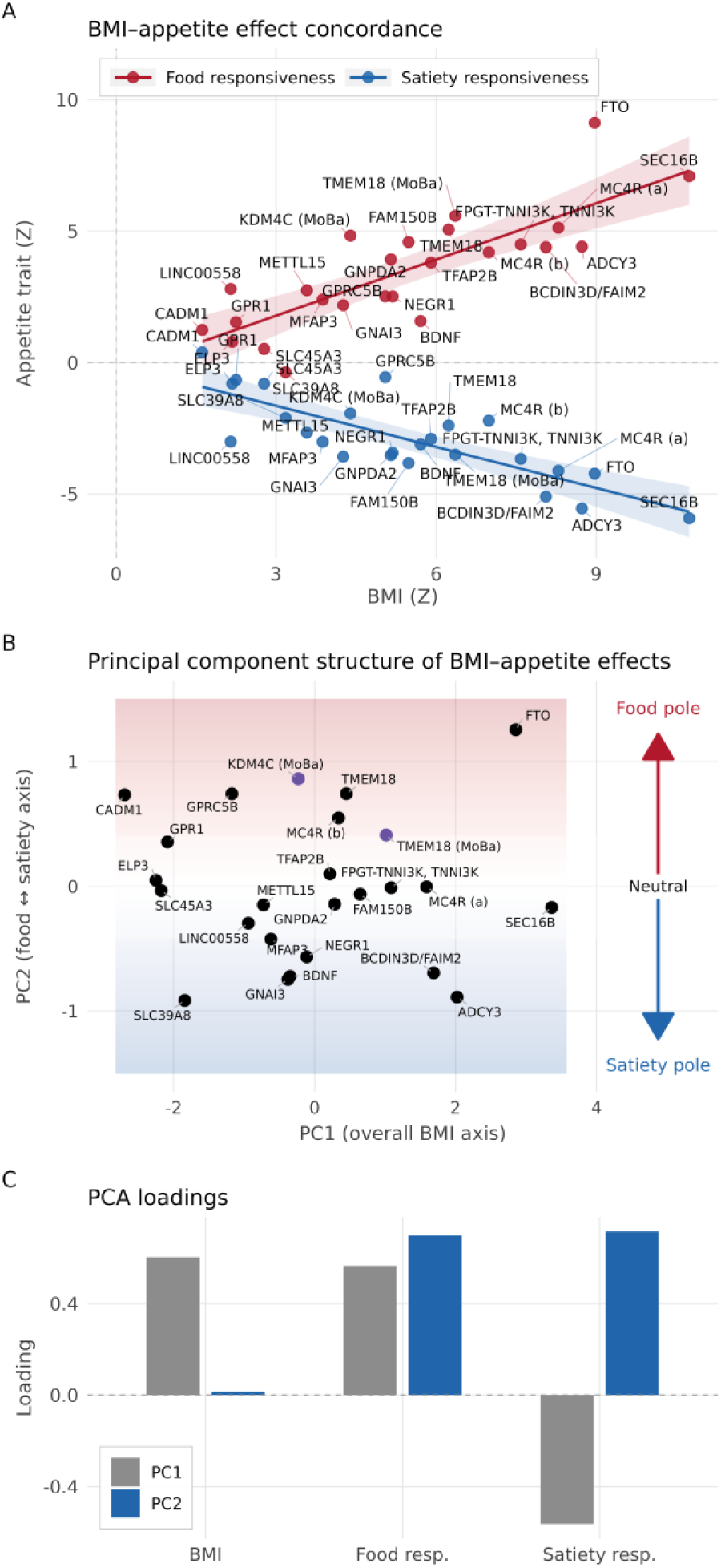
Classification of childhood BMI loci based on their associations with appetitive traits. Genetic effects were examined for 25 loci, comprising 23 established childhood BMI loci from Vogelezang *et al.*, 2020^10^ and two additional BMI at age 8 loci identified in MoBa ( KDM4C and TMEM18). Effects were aligned to the BMI-increasing allele. **(A)** Z-scores for Food Responsiveness ( FR) and Satiety Responsiveness (SR) plotted against the corresponding BMI Z-score at age 8 years in MoBa. **( B)** Principal component (PC) structure of BMI, *FR*, and *SR* effects; PC1 captures adiposity magnitude, PC2 reflects *FR–SR* appetite polarity. **( C)** PCA loadings showing trait contributions to PC1 and PC2 (Table S13). Two independent loci within the *MC4R* region were identified and denoted as *MC4R* ( a) (rs571312) and *MC4R* (b) (rs76227980).

*FTO* shows an outlying *FR* profile among loci with stronger BMI-effects, suggesting its BMI effect is relatively driven by food-approach behaviour, whereas the *ADCY3* and *BCDIN3D/FAIM2* loci align more closely with an *SR* profile. Several additional loci showed intermediate *SR* profiles, including *GNAI3* and *BDNF*. (A third group, near *SEC16B*, *MC4R*(a), *TNNI3K*, and *FAM150B*, clustered near the null in PC2, indicating effects on *FR* and *SR* without clear skew toward either behavioural domain.

### 6. Statistical decomposition of childhood BMI genetic association through appetite traits

Building on the PCA-based variation in how loci relate to *Food Responsiveness* and *Satiety Responsiveness*, we next assessed the extent to which the genetic association with childhood BMI could be statistically decomposed through these appetite traits. We constructed an aggregate Polygenic Risk Score (PRS) of childhood BMI using effect size estimates from Vogelezang *et al*., 2020^10^ and performed regression-based mediation analyses within a counterfactual framework, interpreted here primarily as a statistical decomposition, to quantify the share of the genetic association with BMI captured by *FR* and *SR* (Figure 7A; Table S14 Panel A). Because *FR* and *SR* were observed mediators rather than genetically instrumented exposures, these indirect-effect estimates require strong causal assumptions for a causal interpretation, including no measurement error in the mediator, no unmeasured mediator– outcome confounding and no reverse or bidirectional causality. The PRS was robustly associated with child BMI at age 8. In single-mediator models, 22.1% (95% CI, 20.1–24.1%) of the total PRS–BMI association was statistically captured through *FR* and 10.4% (95% CI, 9.2–11.8%) through *SR*. In a parallel-mediator model including both appetite traits simultaneously, *FR* and *SR* together accounted for 26.7% (95% CI, 24.2–28.9%) of the aggregate genetic association. To assess whether these estimates were sensitive to the use of sum-score appetite domains, we repeated the single-mediator analyses using structural equation models in which *FR* and *SR* were specified as latent factors based on their corresponding CEBQ items. These latent variable models gave similar but slightly larger estimates (*FR*: 24.2% [95% CI, 21.2–27.1%]; *SR*: 13.6% [95% CI, 11.7–15.5%]), suggesting modest increase from sum scoring. (Table S14 Panel A). Sensitivity analyses for unmeasured mediator–outcome confounding indicated that the indirect-effect estimates would require residual mediator–outcome correlations (ρ) of 0.41 for *FR* and −0.26 for *SR* to be attenuated to the null, assuming no empirical mediation effect attenuation due to random measurement error in the mediator.

**Figure 7.**
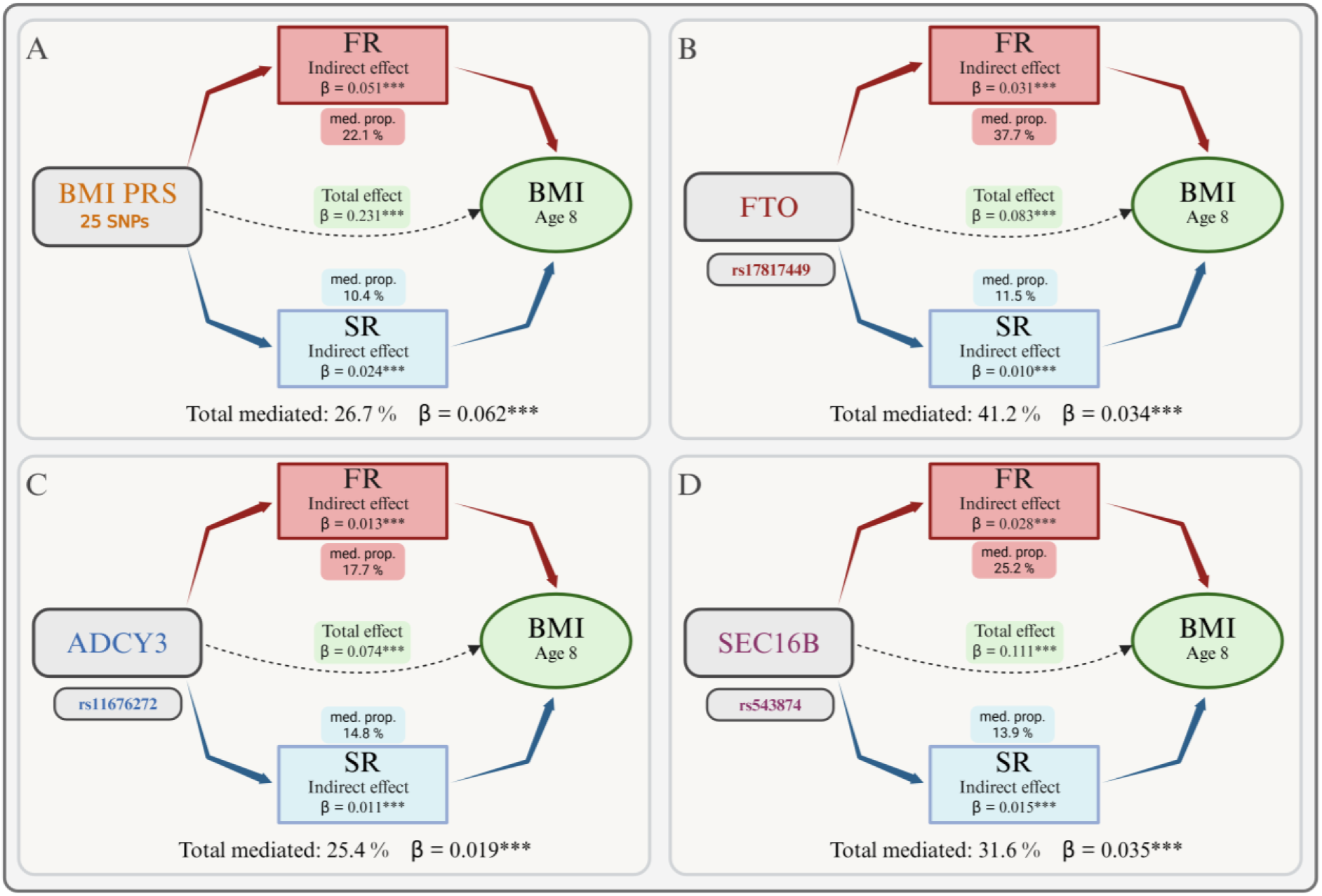
Statistical decomposition of genetic risk for childhood BMI through appetite traits. A) Schematic overview of polygenic risk score (PRS) - level mediation analysis for genetic risk for childhood BMI in relation to BMI at age 8 years through *Food Responsiveness* (*FR*) and *Satiety Responsiveness* ( *SR*). The *FR* and *SR* boxes show single-mediator indirect effects and mediated proportions for each appetite trait separately, whereas the total mediated effect is estimated from the parallel model including *FR* and *SR* simultaneously. B–D) SNP-level mediation models for selected BMI-associated loci: (B) *FTO*, ( C) *ADCY3*, and (D) *SEC16B*. Each panel depicts the total genetic effects on BMI, the trait-specific single-mediator indirect effects through *FR* and SR, and the total mediated effect from the corresponding parallel model.

We next examined SNP-level statistical decomposition for the three loci with the strongest effects on childhood BMI: *FTO*, *ADCY3*, and *SEC16B* (Figure 7B–D). *FTO* showed a clear *FR*-dominant profile, with 37.7% (95% CI, 29.7–48.4%) of the total effect statistically captured through *FR* compared with 11.5% (95% CI, 6.5–17.3%) through *SR* (PFDR < 0.001 for both). This pattern was consistent in latent SEM models of *FR* and remained robust, although attenuated, at age 14 years. *ADCY3* showed a comparatively stronger *SR* contribution than observed for *FTO* or the aggregate PRS pattern, with statistical decomposition through *SR* (14.8% [95% CI, 9.1–21.1%]; P_FDR_ < 0.001) exceeding the PRS-level *SR* estimate, alongside a comparatively smaller *FR* contribution (17.7% [95% CI, 8.3–26.5%]; P_FDR_ < 0.001) relative to *FTO* and the overall PRS pattern. This diverged further at age 14 years, where the *FR* pathway for *ADCY3* attenuated to non-significance, whereas the *SR* pathway remained robust, a pattern replicated in latent SEM model of *SR*. Finally, *SEC16B* aligned with the overall PRS pattern, showing contributions across both pathways at age 8 years (*FR* 25.2% [95% CI, 18.1–32.4%] and *SR* 13.9% [95% CI, 9.3–18.5%], P_FDR_ < 0.001), with convergence at age 14 (*FR* 12.8% [95% CI, 5.5–21.1%] and *SR* 8.3% [95% CI, 3.7–13.2%]), indicating no dominant pathway, a pattern that was consistent in the latent SEM models (Table S15).

As exploratory triangulation, we performed bidirectional two-sample Mendelian randomization (MR) between *FR* and three adiposity measures including childhood BMI^10^, SAC10^18^, and adult BMI^19^ (Table S16). When *FR* was treated as the exposure, genetically instrumented *FR* was associated with all three adiposity measures, although the estimates showed substantial between-instrument heterogeneity (I²=86–98%) and aggregate Steiger tests did not establish FR→adiposity directionality (all P≥0.11). Per-SNP Steiger comparisons revealed marked locus-specific differences. The *FTO*-region variant rs9937053 was retained for childhood BMI and body size at age 10, whereas the *ADCY3*-region variant rs11676272 showed statistically supported adiposity orientation for both traits (P=0.0022 and 0.025). After per-SNP Steiger filtering, the FR→childhood BMI association persisted and heterogeneity was no longer evident (Table S16).

When adiposity traits were treated as exposures, childhood BMI, body size at age 10 and adult BMI were each associated with greater *FR*, with aggregate Steiger tests supporting adiposity→FR directionality in all three analyses. However, directional pleiotropy for body size at age 10 and adult BMI remained evident. Together, these analyses support a shared and potentially bidirectional appetite–adiposity genetic architecture, while the locus-specific differences in Steiger orientation complement the heterogeneous appetite profiles observed in the GWAS and mediation analyses.

### 7. Nature or nurture: the role of direct and indirect genetic effects on eating behaviour

In addition to direct genetic effects, a child’s appetite and BMI may be influenced by the family environment shaped by parental genetic liability, often referred to as “genetic nurture”. The unique design of MoBa, in which both parents and children have been genotyped, allowed us to use genetic instruments to address whether the parentś own BMI influences their perception of their own child’s eating behaviour and how this relates to the inherited component shared by offspring and parents. To distinguish direct genetic effects from indirect parental influences, we applied a trio-based polygenic risk score (PRS) decomposition in parent–offspring full trios (N= 48645). We constructed a ∼900-SNP BMI PRS based on the adult BMI GWAS by Yengo *et al.* (2018)^19^ as an instrument for parental BMI. The PRS showed strong associations with BMI in both mothers and fathers (mothers: β = 1.03, R² = 5.6%; fathers: β = 0.79, R² = 5.6%). By phasing the children’s alleles to infer whether a parental allele was transmitted to the child or not, we then computed this PRS separately for maternal transmitted (MT), paternal transmitted (PT), maternal non-transmitted (MnT), and paternal non-transmitted (PnT) alleles within each trio. Transmitted scores estimate inherited child genetic effects while reducing bias from parental genotype and family-level confounding, whereas non-transmitted scores capture parental genetic liability not inherited by the child and therefore provide a proxy for indirect parental influences.

The child’s inherited genetic liability, captured by the MT and PT scores, was robustly associated with their longitudinal BMI and increased from infancy through age 14 (Figure 8A) at similar levels for both the maternal and paternal transmitted score. This is in line with our previous work showing increasing predictive performance throughout childhood for adult-based BMI-PRSs (Helgeland *et al*., 2019^20^; 2022^11^). The non-transmitted parental polygenic scores (MnT and PnT), indexing parental genetic liability of increased BMI *not* inherited by the child, showed near-zero association with childhood BMI across all postnatal timepoints. A transient maternal effect was observed at birth, likely reflecting intrauterine influences of the mother’s BMI, but this attenuated rapidly and converged with paternal estimates thereafter.

**Figure 8.**
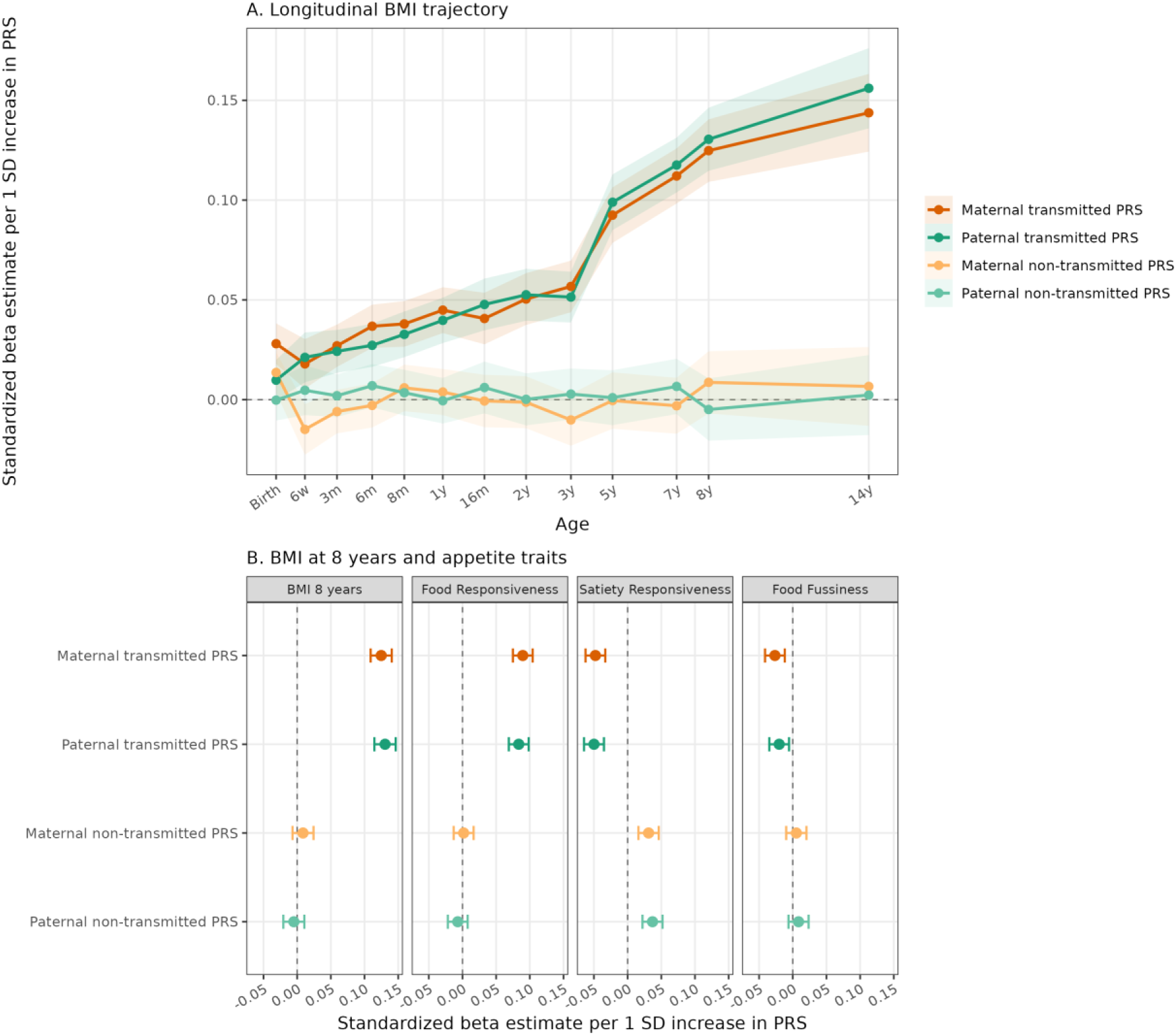
Direct genetic and environmental (genetic nurture) effects on child BMI and appetite. A) Longitudinal BMI trajectory: Maternal and paternal transmitted adult BMI polygenic risk scores (MT and PT) versus non-transmitted scores (MnT and PnT) on child BMI z-scores from birth to 14 years. B) Polygenic effects at age 8: Forest plot comparing the independent effects of the same four PRS components on child BMI age 8, Food Responsiveness, Satiety Responsiveness, and Food Fussiness. Points represent standardized beta estimates (per 1 SD increase) from joint linear regression models including all four PRSs simultaneously. Shaded bands (A) and error bars ( B) indicate 95% confidence intervals. All models are adjusted for child sex and 10 genetic principal components, utilizing cluster -robust standard errors to account for sibling clustering.

Children’s BMI and *FR* at 8 years were both driven mainly by transmitted genetic scores, showing a strong positive association with MT and PT adult-BMI-PRSs, but no statistically significant association with either the maternal or paternal non-transmitted scores. We found no evidence that parental genetic liability influenced ratings of *FF*. However, *SR* showed evidence of both direct and indirect genetic influences: the child’s transmitted BMI-increasing alleles were associated with lower *SR*, whereas parental non-transmitted adult BMI PRS (MnT and PnT) were associated with higher *SR* (P < 0.001; Figure 8B; Table S14, Panel B). Because MnT and PnT are independent of the child’s genotype (and were themselves essentially uncorrelated, ρ ≈ 0, indicating no assortative mating), this pattern may reflect indirect parental effects on the feeding environment, subjective reporting bias where the parents’ own BMI alters their perception of the child’s *SR*, or both, and in the opposite direction to the effects of transmitted alleles.

Notably, adjusting the mediator-to-BMI path in the PRS mediation models for both MnT and PnT only changed the indirect effects of the child’s PRS via *FR* and *SR* by less than 1% (Table S14, Panel A). Together with the latent variable results, this indicates that the appetite-related mediation pathways primarily reflect the child’s own inherited genetic effects rather than parental indirect genetic influences.

## Discussion

Using data from the large pregnancy-based MoBa cohort, this study provides genome-wide evidence that *Food Responsiveness* (FR) and *Satiety Responsiveness* (SR) as captured by parents’ report, are under considerable genetic influence and mainly driven by the child’s own genetic makeup rather than parental indirect effects. *FR* and *SR* share substantial genetic architecture with BMI from early childhood through adolescence, together statistically capturing over a quarter of the aggregate genetic association with BMI at age 8 years. Food Fussiness, despite being the most heritable trait, showed only weak genetic overlap with BMI, and none of the other domains yielded genome-wide significant signals. Locus-specific analyses identify mechanistically distinct pathways of appetite regulation, linking common variants to paediatric adiposity, consistent with the Behavioural Susceptibility Theory (Llewellyn *et al*., 2023^21^).

The genome-wide significant loci identified for appetite largely overlap established childhood BMI loci, consistent with the possibility that a substantial proportion of adiposity-associated genetic risk operates through appetite-regulatory neuronal and hypothalamic circuits (Krashes *et al*., 2016^22^). Notably, some loci differed systematically in their associations with specific appetitive domains, suggesting heterogeneity in the behavioural pathways linking genetic risk to adiposity. The *FTO* locus showed a disproportionately strong association with *FR*, consistent with prior evidence indicating that its association with appetite is not fully explained by BMI and may be reflected in increased *Food Responsiveness*, captured by parents through the item “*If allowed, my child would eat too much”*. In children aged 10-11 in ALSPAC, risk allele carriers consumed significantly more total energy and fat even after adjustment for BMI (Timpson *et al*., 2008^14^). Similarly, Velders *et al.* (2012)^23^ demonstrated in a population-based cohort that the *FTO* risk allele was already associated with elevated *FR* at age 4, when no clear association with BMI was yet detectable in that cohort. One possible interpretation is that this apparent BMI-null window may not reflect biological inactivity, but rather two opposing effects that may partially offset one another during early childhood: an increase in *FR* on one hand, and the known negative early-life effect of *FTO* on BMI on the other (Helgeland *et* al., 2019^20^). This interpretation is supported by mediation analyses (Emond *et al*., 2017^24^) and by neuroimaging data showing persistent postprandial activation of reward-related corticolimbic regions including the ventral striatum and amygdala in high-risk allele carriers, which are associated with increased energy intake (Melhorn *et al*., 2018^25^). By contrast, *ADCY3* showed a relatively stronger association with *SR*, consistent with its role as the molecular effector of *MC4R* anorexigenic signalling hypothalamic neurons (Siljee *et al*., 2018^16^). Rare loss-of-function *ADCY3* mutations cause severe hyperphagia-driven obesity in humans (Saeed *et al*., 2018^26^; Özcabı *et al*., 2025^27^), and common variants reduce hypothalamic expression (Stergiakouli *et al*., 2014^28^), collectively pointing to impaired satiety signalling. Taken together, these findings could be interpreted as supporting a pathway-specific model, where the *FTO* variant is more related to food cue reactivity and reward processing, whereas *MC4R*–*ADCY3* signalling may more directly regulate hypothalamic satiety, potentially informing targeted preventive strategies.

Among other loci with the strongest effects on childhood BMI, we note that signals near *TMEM18* appeared more associated with *FR* than *SR*, while the opposite patterns were seen for *FAIM2* – a gene recently proposed to affect obesity through altered feeding behaviour (Littleton *et al*., 2024^29^). The *TMEM18* results are consistent with leptin-sensitive expression in the hypothalamic paraventricular nucleus driving food intake in animal models (Larder *et al*., 2017^30^), though the relatively modest behavioural signal likely reflects a substantial contribution from appetite-independent adipogenic pathways (Landgraf *et al*., 2020^31^). In contrast, the *SEC16B* locus that confers the largest BMI-effect at age 8 years showed less appetite pathway specificity. Intestinal knockout mice are protected from diet-induced obesity through impaired chylomicron lipidation rather than reduced food intake (Shi *et al*., 2023^32^), suggesting that its association with appetitive traits in our data likely reflects indirect metabolic feedback rather than a primary effect on appetite regulation. Together, these results illustrate how eating behaviour GWASs may provide mechanistic insights and help decompose polygenic BMI risk into central or peripheral modes of action.

*Food Fussiness* is of particular interest, as associations at both *SIX3* and the *17q21.31 inversion polymorphism* remained unchanged after BMI adjustment, supporting an obesity-independent link with picky eating. The 17q21.31 region comprises a ∼900 kb inversion polymorphism with two major haplotypes, the ancestral (H1) and inverted (H2), as well as multiple sub-haplotypes that vary substantially in frequency across populations (Stefansson *et al*., 2005^33^; Pedicone *et al*., 2024^34^), offering opportunities for future studies to further explore the underlying genetic architecture. At the *SIX3* locus, the lead SNP rs504675 has previously been associated with food-liking traits, particularly preferences for bitter, spicy, salty, fatty, and other strong or acquired flavours (May-Wilson *et al*., 2022^35^). In contrast, prior fine-mapping efforts at the broader *SIX2/SIX3* region have focused on glycaemic traits, but the reported lead variant (rs12712928) appears to be independent of the *Fussiness* signal observed here (Spracklen *et al*., 2018^36^).

At the aggregate level, the childhood-based BMI polygenic risk score showed stronger statistical decomposition through *FR* than through *SR*, suggesting that food cue reactivity captures a larger share of the PRS–BMI association than *Satiety Responsiveness*. The magnitude observed in our statistical mediation, over a quarter of the PRS-BMI association at age 8 years captured by currently identified childhood BMI variants, exceeds prior estimates: longitudinal evidence found that overeating mediated 11–18% of the genetic association with BMI across childhood, attenuating over time, with fussy eating showing no contribution (Goulet *et al*., 2025^37^), and broader obesogenic appetite profiles explain around one-fifth of the polygenic risk–BMI association in paediatric samples (Renier *et al*., 2024^38^). Parallel mediation evidence from adult UK cohorts similarly implicates food approach behaviours as the dominant eating behaviour mediators of BMI polygenic risk (Begum *et al*., 2023^39^), suggesting that the prominence of *FR* is consistent across the life course. The locus-level pathway specificity we observe, with *FTO* showing an *FR*-dominant profile and *ADCY3* showing a relative shift toward *SR*, maps onto this aggregate pattern and is consistent with prior mediation evidence in children (Emond *et al*., 2017^24^; Llewellyn *et al*., 2014^40^), supporting the interpretation that the overall dominance of *FR* may partly reflect the aggregate contribution of *FTO*-like loci rather than a uniform mechanism across all adiposity variants.

Phenotypic correlations between child and parental BMI, and between *FR* and parental BMI, can be due to contributions from both genetic transmission and shared family environments. However, evidence from family-based studies regarding the role of parental genetic nurture has been inconsistent, with reports ranging from negligible effects (Schnurr *et al*., 2020^41^) to substantial maternal indirect genetic influences (Wright *et al*., 2025^42^). Thanks to a large sample size and a trio-based decomposition of genetic transmission, our analyses provide more precise estimates of transmitted and non-transmitted parental genetic associations and help clarify these conflicting findings. We show that transmitted alleles (direct genetic effect) capture the predominant inherited genetic component of childhood BMI and *FR*, whereas non-transmitted parental BMI-associated alleles (genetic nurture) show minimal impact across early and mid-childhood. A distinct pattern emerged for *SR*, where non-transmitted parental BMI-increasing alleles were associated with higher parent-reported *Satiety Responsiveness* of their children, despite minimal effects on the child’s BMI. This dissociation suggests that parent-reported *SR* (but not *FR*) may partly reflect parental perception rather than the child’s intrinsic biology, providing insights in agreement with previous studies that have suggested that parental genetic obesity risk shapes appetite perception and reporting (Jansen *et al*., 2023^43^). Although the lack of correlation between MnT and PnT scores reduces concern about assortative mating as the main explanation, non-transmitted PRS associations should still be interpreted cautiously, as they may also reflect population stratification or residual family-level confounding.

Item-level genetic signals closely paralleled domain-level estimates for *FR*, and more modestly for *SR*, but were weaker and less consistent across other domains. This suggests that selected items may approximate domain-level genetic signal for specific traits, particularly *FR*, and could be considered as pragmatic proxies in large-scale settings where it may not be feasible to include a more comprehensive set of questions. Emotional eating showed notably weaker heritability, consistent with twin evidence for dominant shared environmental influence in early childhood (Herle *et al*., 2017^44^; Madhavan *et al*., 2025^45^), while *Food Fussiness* showed the highest SNP-heritability (Nas *et al*., 2025^46^), yet a weak BMI association, pointing to a biologically distinct architecture driven by genes largely separate from those shaping adiposity.

There are some limitations of the study. Although the mediation findings were robust across sensitivity analyses and temporal extension to adolescent BMI, the possibility of residual unmeasured confounding in the appetite–BMI pathway means estimates should be interpreted with caution rather than as reflecting a fully isolated causal mechanism. In particular, while Behavioural Susceptibility Theory supports appetite traits as plausible behavioural pathways linking genetic susceptibility to adiposity, higher BMI may also influence later appetite, feeding dynamics, or parent-reported eating behaviour across development. Consistent with this possibility, bidirectional MR showed more consistent Steiger directionality from adiposity to FR, although differences in instrument strength, phenotype measurement and evidence of pleiotropy prevent definitive inference about causal direction. Parent-reported appetite, while well-validated and widely used, may not fully capture the child’s intrinsic appetitive biology, particularly for *SR*, where parental perception is itself shaped by parental characteristics as demonstrated in this study. However, it is reassuring that the effects of parental non-transmitted alleles on SR were directionally opposite to those of transmitted alleles, making positive confounding by parental characteristics or reporting tendencies less likely. Future work incorporating observational or neuroimaging measures of appetite alongside genetic data would strengthen causal inference. These findings await replication in independent cohorts with GWAS and parent-reported eating behaviour data. It should also be noted that BMI was parent-reported at age 8 and self-reported at age 14, introducing distinct sources of measurement bias. Parent-reported BMI may attenuate associations with health outcomes (Shields *et al*., 2011^47^), suggesting our primary age-8 mediation estimates may be conservative, while self-reported BMI for age 14 years may be less susceptible to bias (Rios-Leyvraz *et al*., 2022^48^). Although both timepoints are subject to reporting error, the underlying bias structure differs between parent-reported BMI at age 8 and self-reported BMI at age 14, making it less likely that the observed pattern is driven solely by a shared reporting artefact. The predominantly European sample additionally limits generalisability. Nonetheless, the stronger statistical contribution of *FR* suggests that children with high genetic liability for BMI who also carry heightened *FR* may represent a priority population for responsive feeding interventions targeting food cue exposure and feeding environment regulation. Genetic risk stratification as a tool for identifying such children warrants prospective evaluation in intervention studies before clinical implementation can be considered.

## Conclusion

This study demonstrates that childhood appetitive traits are heritable and play an important role in the development of obesity early in life. Genome-wide analyses show that these traits share genetic factors with BMI, while also exhibiting distinct genetic signals. We further show that obesity-associated genetic variants are linked to different dimensions of appetite, with some operating primarily through heightened responsiveness to food cues and others through variation in satiety-related processes. Using a parent-offspring trio design, we also show that parent-reported appetite is predominantly explained by the child’s own genotype, with little influence from parental indirect genetic effects. Together, these findings advance our understanding of how genetic risk for obesity is expressed through eating behaviour early in life and highlight appetite regulation as a potential target for early prevention of obesity.

## Methods

### Study population

Data were drawn from the Norwegian Mother, Father and Child Cohort Study (MoBa), a population-based pregnancy cohort conducted by the Norwegian Institute of Public Health (NIPH) (Magnus *et al*., 2016^49^; Brandlistuen *et al*., 2025^50^). The study includes approximately 114,500 children, 95,200 mothers, and 75,000 fathers, recruited from 50 hospitals across Norway between 1999 and 2008. Around 41% of invited pregnancies participated. The establishment of MoBa was approved by the Norwegian Data Protection Authority, and the study is regulated under the Norwegian Health Registry Act. The present analyses were approved by the Regional Committees for Medical and Health Research Ethics (REK: #2012/67).

### Child Eating Behaviour Questionnaire (CEBQ)

Appetitive traits were measured using a short-form version of the Child Eating Behaviour Questionnaire (CEBQ), a validated parent-reported tool assessing children’s eating styles and appetite regulation tendencies (Wardle *et al*., 2001^3^). The current instrument in MoBa comprised 18 items covering six behavioural domains: *Food Responsiveness (FR), Satiety Responsiveness (SR), Food Fussiness (FF), Slowness in Eating (SiE), Emotional Undereating (EU),* and *Emotional Overeating (EO)*. Parents rated 18 items on a five-point Likert scale (1 = Never, 5 = Always). The three positively worded FF items were reverse scored so that higher domain scores reflected greater expression of the corresponding trait.

Psychometric structure was evaluated in 28,580 children, split by family into independent EFA (N = 14,292) and CFA (N = 14,288) samples. EFA used polychoric correlations, minimum-residual extraction and oblimin rotation. parallel analysis and comparison of one- to eight-factor solutions supported retention of the six-domain structure (Table S2). The prespecified six-factor model was then tested by WLSMV CFA for ordinal indicators. Standardized CFA loadings ranged from 0.56–0.98 and Cronbach’s α from 0.70–0.92; model-fit statistics are reported in Table S2 (Figure 1; Table S2).

### BMI and anthropometric phenotypes

Child height and weight were measured by trained health personnel at routine examinations at birth and 6 weeks and reported by parents at 3, 6, and 8 months, and at 1, 1.5, 2, 3, 5, 7, and 8 years; self-reported data at age 14 years were available for a subset. BMI (kg/m²) was standardized within age and sex using the LMS method implemented via the generalised additive model for location, scale and shape (GAMLSS, v5.1-7), as described in Helgeland *et al*. (2022). Extreme values (|z| > 5) were winsorized. Parental BMI was derived from pre-pregnancy self-report for mothers and enrolment-time data for fathers.

### Genotyping, quality control, and ancestry

Analyses were based on imputed genotypes from the MoBa cohort and were restricted to individuals of European ancestry with complete phenotype and covariate data. Genotyping data were generated using various genotyping platforms by multiple Norwegian research groups and are currently curated by the Norwegian Institute of Public Health (NIPH). The genotyping batches used in this study are documented and accessible via the NIPH repository (github.com/folkehelseinstituttet/mobagen); version 1.5 of the genotype dataset was utilized for all analyses. Quality control procedures were carried out by the PsychGen team, with full details available in their published pipeline description (Corfield *et al.*, 2024^51^) and associated codebase (github.com/psychgen/MoBaPsychGen-QC-pipeline). Related children were retained in the GWAS analyses, with relatedness accounted for by REGENIE’s whole-genome regression framework.

### Genome-wide association analyses

GWAS were performed for each of the six domain scores and 18 individual items (24 traits in total) using REGENIE v3.2 (Mbatchou *et al*., 2021^52^), available via Bioconda (Grüning *et al*., 2018^53^), and executed via a Snakemake-based workflow (Mölder *et al*., 2021^54^) on the HUNT Cloud computing infrastructure. REGENIE implements a two-step whole-genome regression approach that efficiently accounts for sample relatedness and population stratification.

In Step 1, a ridge regression model was fitted on directly genotyped variants passing quality filters (MAF > 0.01, call rate > 0.98, Hardy–Weinberg equilibrium P > 1×10⁻⁶) to generate polygenic predictions. In Step 2, imputed SNP dosages (INFO > 0.8) were tested for association with each phenotype. All models were adjusted for child sex, genotyping batch, and the first ten genetic principal components. Genomic inflation factors (λ < 1.05) for all traits indicated minimal population stratification bias. Genome-wide significance was defined as P < 5 × 10⁻⁸. The effective number of independent tests was estimated separately for domains (Meff = 5.7) and items (Meff = 16.8) using the approach of Nyholt (2004)^55^ and Nyholt-adjusted P values are provided in Table S10.

To classify established childhood BMI loci by their appetitive mechanism, we retrieved Z-scores (Beta/SE) for BMI at age 8, *Food Responsiveness*, and *Satiety Responsiveness* from MoBa for the 23 lead SNPs from Vogelezang *et al*. (2020)^10^ plus 2 SNPs from MoBa, aligning all effects to the BMI-increasing allele, and applied PCA to partition loci by their food-approach versus satiety association profiles (Table S13).

### SNP-based heritability and genetic correlations

SNP-based heritability (h²) and pairwise genetic correlations (r_g_) were estimated using LD Score Regression (LDSC) (Bulik-Sullivan *et al*., 2015^56^). GWAS summary statistics were pre-processed using munge_sumstats.py, retaining HapMap3 SNPs (International HapMap 3 Consortium *et al*., 2010^57^) with MAF > 0.01. The 1000 Genomes European-ancestry LD reference panel was used throughout. We estimated: (i) SNP heritability for each of the 24 appetitive traits; (ii) pairwise genetic correlations among all 24 traits; and (iii) genetic correlations between each CEBQ domain and standardised BMI at 13 timepoints from birth to 14 years, comparative body size at age 10 (SAC10, UK Biobank), and adult BMI (GIANT/UK Biobank meta-analysis). Genetic correlations with a broad panel of external GWAS traits including anthropometric, metabolic, dietary, hormonal, cardiovascular, and psychological phenotypes were estimated. Statistical significance across all LDSC tests was assessed using false discovery rate (FDR) correction (Benjamini–Hochberg).

### Locus definition and annotation

Independent association signals were first defined using conditional and joint multiple-SNP analysis (COJO) implemented in Genome-wide Complex Trait Analysis (GCTA) (Yang *et al*., 2012^58^) on the Regenie-derived GWAS summary statistics. COJO identifies conditionally independent variants by jointly modelling all SNPs within a ±500 kb region, using the MoBa genotype data as the LD reference. At loci with more than one conditionally independent variant, the primary signal is reported with its marginal association statistics and any secondary signal with its COJO joint estimate; secondary signals were retained if the joint P value passed the genome-wide threshold.

Because several of the domain-level traits (e.g., *FR*) and their individual items (e.g., “would eat too much,” “would always have food”) are expected to capture overlapping biological signals, we next applied colocalization analysis using the coloc R package (Giambartolomei *et al*., 2014^59^). For every pair of COJO lead variants within the same 500 kb window across related traits, coloc.abf() was used to estimate the posterior probability of sharing a common causal variant (PP₄). Loci with PP₄ > 0.8 were considered colocalized, and only the most significant lead SNP (lowest p-value) from each colocalized group was retained. This two-step procedure, COJO for statistical independence within each trait and coloc for biological overlap across correlated traits, ensured that each genomic locus was counted once, yielding a non-redundant set of appetite-associated loci for downstream annotation and visualization.

### Mediation and sensitivity analyses

We assessed whether genetic susceptibility to adiposity was statistically mediated through *Food Responsiveness* (*FR*) and *Satiety Responsiveness* (*SR*) using both polygenic and single-locus approaches.

Genetic effects were modelled at both the single-locus and polygenic level, with the exposure defined as either SNP dosage (aligned to the BMI-increasing allele for *FTO*, *ADCY3*, and *SEC16B*) or a standardized childhood BMI polygenic risk score (PRS; Vogelezang *et al*., 2020^10^). *FR* and *SR* were examined as mediators, and BMI z-score at age 8 years as the primary outcome. For the primary observed-score mediation analysis *FR* and *SR* were modelled using standardized CEBQ domain scores. We fitted single-mediator models, in which *FR* and *SR* were examined separately, and a parallel two-mediator model, in which *FR* and *SR* were included simultaneously to account for their covariance. In the single-mediator models, the indirect effect was estimated as the product of the genetic predictor–mediator association and the mediator–BMI association. In the parallel model, separate indirect effects were estimated for *FR* and *SR* conditional on the other appetite trait, and the total indirect effect was defined as the sum of the *FR* and *SR* indirect effects. Direct, indirect, total, and proportion-mediated estimates were obtained using regression-based product-of-coefficients models.

All models were adjusted for child sex and the first ten genetic principal components, with confidence intervals obtained using bias-corrected and accelerated bootstrap resampling (1,000 iterations). To assess robustness to measurement error in parent-reported appetite, we additionally re-estimated mediation pathways using latent representations of *FR* and *SR* based on their corresponding CEBQ items in sensitivity analysis. Analyses using BMI at age 14 years were performed as a temporal extension. Robustness was further evaluated by quantifying the degree of unmeasured mediator–outcome confounding required to attenuate indirect effects to zero using sensitivity analysis of the residual correlation parameter ρ (Imai *et al*., 2010^60^).

The mediation analyses rely on assumptions that the genetic predictor, mediator, and outcome are correctly ordered; that there is no unmeasured confounding of the genetic predictor– mediator, genetic predictor–outcome, or mediator–outcome associations; that there is no mediator–outcome confounder affected by the genetic predictor; and that reverse or bidirectional relationships between appetite and BMI do not fully explain the observed indirect effects. Although genetic predictors reduce some sources of confounding, horizontal pleiotropy, residual population structure, assortative mating, parental genetic effects, and measurement error in parent-reported appetite or BMI could still bias estimates. Given the concurrent measurement of appetite and BMI at age 8 years, mediation estimates are therefore interpreted as statistical decompositions of the genetic association with BMI rather than definitive causal effects.

For locus-level mediation analyses, multiple testing was controlled using the Benjamini– Hochberg false discovery rate (FDR) across the prespecified primary tests, comprising the indirect and total effects across loci, mediators and BMI outcomes

### Mendelian Randomization

As exploratory triangulation, we performed bidirectional two-sample Mendelian randomization (MR) between standardized *FR* and childhood BMI, comparative body size at age 10, and adult BMI. For *FR*→adiposity analyses, instruments were eight LD-independent variants representing loci identified across the FR domain- and item-level GWAS, with SNP– *FR* associations obtained from the standardized FR domain GWAS. For adiposity→FR analyses, genome-wide significant variants (P≤5×10⁻⁸) were selected independently from each adiposity GWAS. Variants were LD-clumped against the 1000 Genomes European reference panel (r²<0.001 within 500 kb) and harmonised in TwoSampleMR (action=2). Instrument strength was assessed using F statistics. Inverse-variance weighted (IVW) MR was the primary estimator, with weighted-median and MR-Egger analyses as sensitivity estimators; heterogeneity, directional pleiotropy and influential variants were assessed using Cochran’s Q, the MR-Egger intercept and leave-one-out analyses, respectively. Directionality was evaluated using aggregate and per-variant Steiger tests, and MR was repeated after excluding variants explaining more variance in the outcome than the exposure as a sensitivity analysis (Table S16).

### Genetic nurture and trio-based PRS decomposition

To disentangle direct genetic effects from indirect parental influences, we decomposed an adult BMI PRS into transmitted and non-transmitted maternal and paternal components using trio data. PRSs were constructed by aggregating weighted contributions from maternal transmitted (MT), maternal non-transmitted (MnT), paternal transmitted (PT), and paternal non-transmitted (PnT) alleles.

Associations with child BMI across the life course, as well as *FR* and *SR*, were tested in joint models including all four components simultaneously, with family-clustered standard errors to account for sibling structure. Non-transmitted PRSs were further incorporated into mediation models to evaluate the extent to which parental genetic liability influenced estimated behavioural pathways. Assortative mating was assessed via correlations between maternal and paternal PRS components, which were near zero, suggesting minimal influence on indirect genetic effect estimates.

### Software and reproducibility

All analyses were conducted in R (v4.2.2), Python (v3.10), and REGENIE (v3.2). Workflow orchestration and dependency management were handled using Snakemake (v7.0+).

## Ethics declarations

MoBa: Informed consent was obtained from all study participants. The administrative board of the Norwegian Mother, Father and Child Cohort Study led by the Norwegian Institute of Public Health approved the study protocol. The establishment of MoBa and initial data collection was based on a license from the Norwegian Data Protection Agency and approval from The Regional Committee for Medical Research Ethics. The MoBa cohort is currently regulated by the Norwegian Health Registry Act. The study was approved by The Regional Committee for Medical Research Ethics (#2012/67).

## Data Availability

Individual-level data from the Norwegian Mother, Father and Child Cohort Study (MoBa) are not publicly available due to participant consent and data protection requirements. Researchers may apply for access to MoBa data, including genetic data, through the Norwegian Institute of Public Health, subject to the relevant ethical and data-access approvals. External GWAS summary statistics used in this study are available from the sources cited in the manuscript. Summary-level results generated in the present study are reported in the manuscript and Supplementary Information.

https://www.fhi.no/en/ch/studies/moba/for-forskere-artikler/research-and-data-access/

## Acknowledgements

We thank the Norwegian Institute of Public Health (NIPH) for generating high-quality genomic data, we thank the Norwegian Institute of Public Health (NIPH), the HARVEST collaboration, the NORMENT Centre at the University of Oslo, the Center for Diabetes Research at the University of Bergen, deCODE Genetics, the Research Council of Norway, the SouthEastern and Western Norway Regional Health Authorities, the ERC AdG, Stiftelsen KG Jebsen, the Trond Mohn Foundation, and the Novo Nordisk Foundation. We thank the MoBaPsychGen team, led by Elizabeth Corfield, for providing quality controlled genotype data, supported by funding from the South-Eastern Norway Regional Health Authority [2021045; 2020022; 2022083; 2018058].

We are grateful to all the families in Norway who are taking part in the ongoing MoBa cohort study. All analyses in the MoBa cohort were performed using digital labs in HUNT Cloud at the Norwegian University of Science and Technology, Trondheim, Norway. We are grateful for outstanding support from the HUNT Cloud community.

