## Supplementary Figures S1-S4 for "Genetic and behavioural architecture of childhood eating behaviour and links to obesity risk"

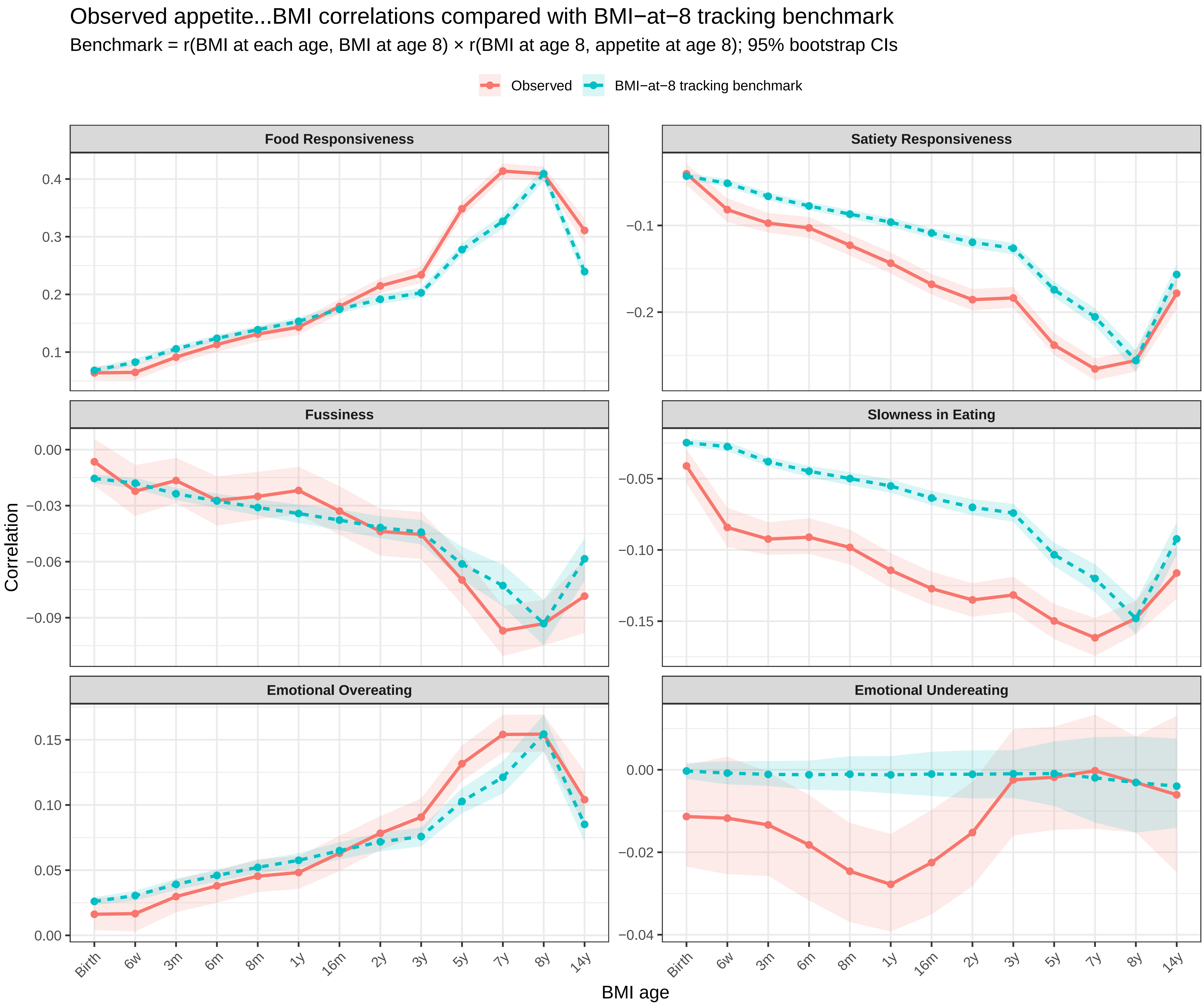


Figure S1. Observed appetite–BMI correlations compared with a BMI-at-8 tracking benchmark. The observed correlation is the Pearson correlation between each appetite domain at age 8 years and BMI at each age. The BMI-at-8 tracking benchmark was calculated as r(BMI at each age, BMI at age 8) × r(BMI at age 8, appetite at age 8), representing the correlation anticipated if the association at another age reflected BMI tracking through age 8 alone. Shaded areas indicate 95% bootstrap confidence intervals.

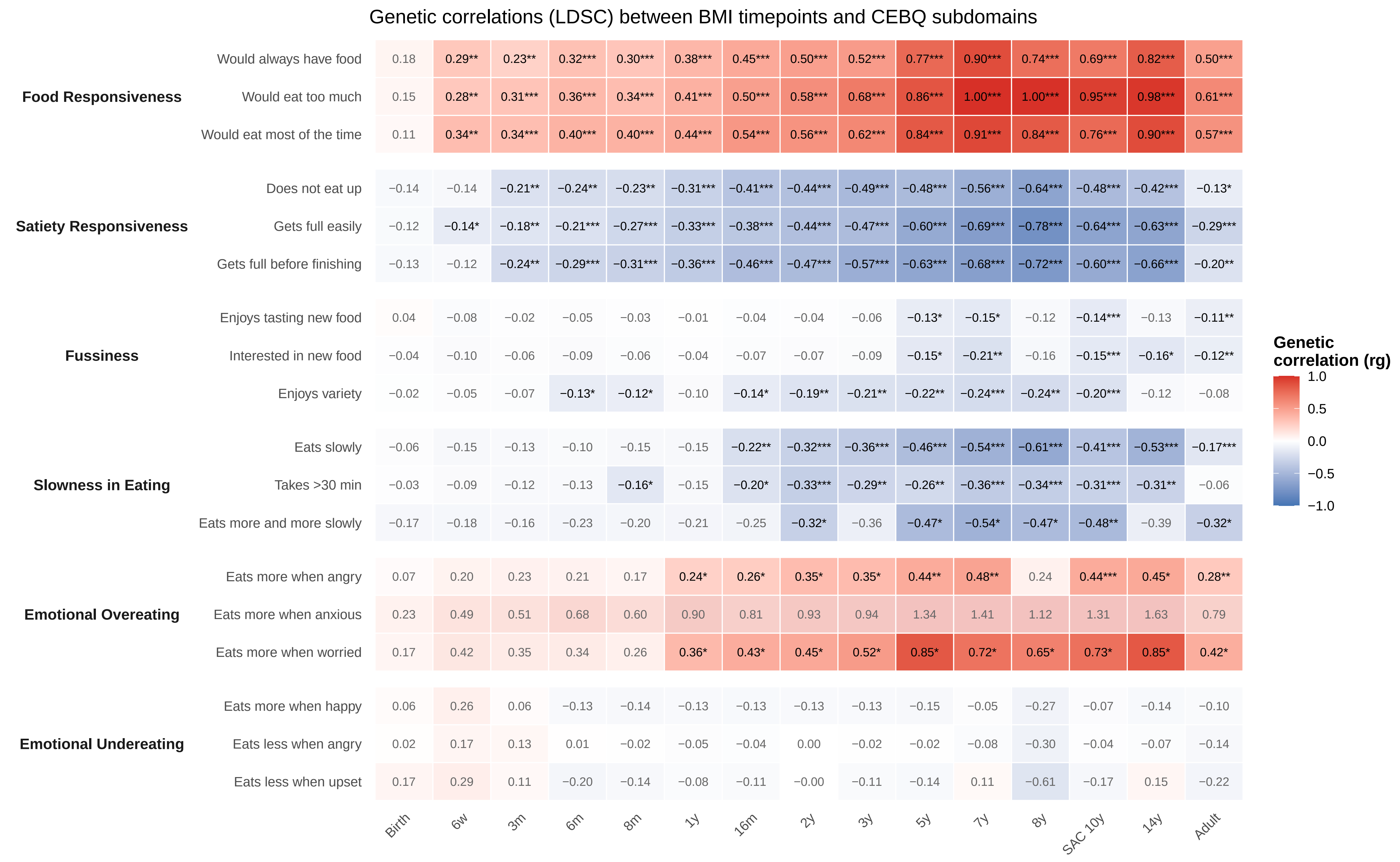


Figure S2. Genetic correlations between BMI across the life course and CEBQ subdomain items.
Heatmap showing LD Score Regression (LDSC)–based genetic correlations (rg) between BMI measured from birth through adulthood and individual items from the Child Eating Behaviour Questionnaire (CEBQ) at age 8 years. Items are grouped by domain: Food Responsiveness, Satiety Responsiveness, Food Fussiness, Slowness in Eating, Emotional Overeating, and Emotional Undereating. Asterisks denote statistical significance (*P < 0.05, **P < 0.01, ***P < 0.001).

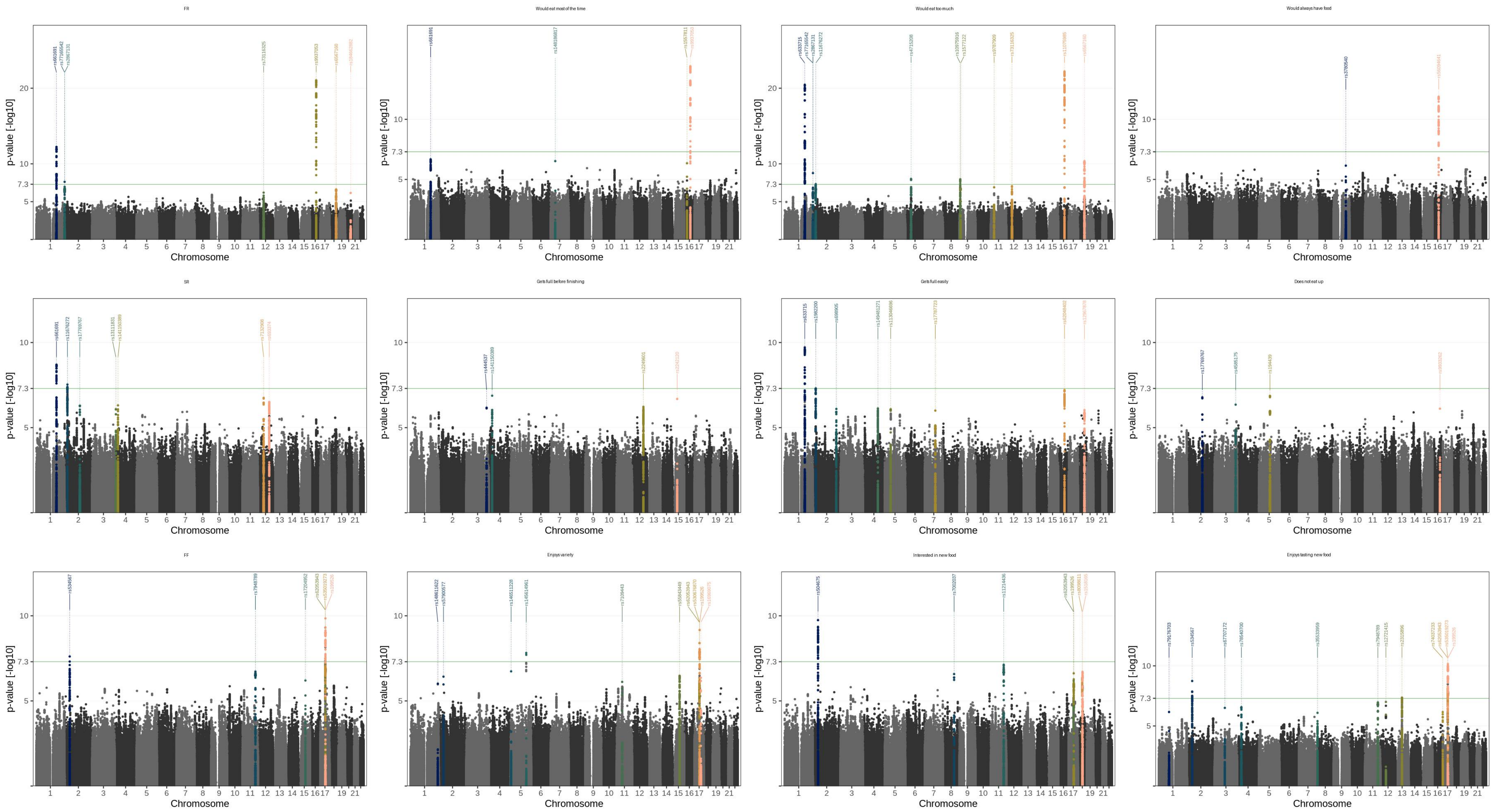


Figure S3. Manhattan plots for CEBQ appetite domains and items. Item codes refer to question texts listed in Table S1; dashed lines mark genome-wide significance ($p < 5\times10^{-8}$).

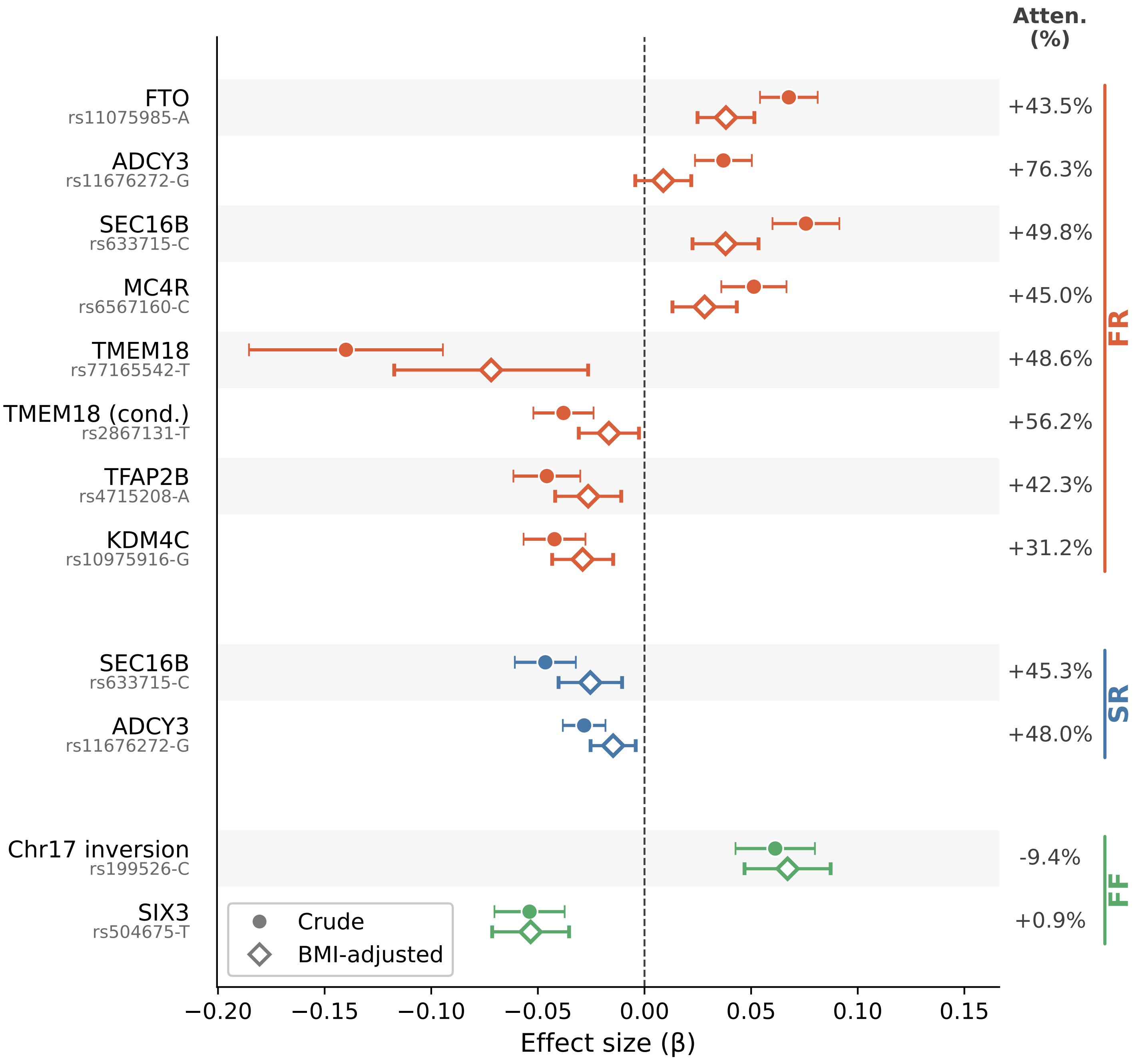


Figure S4. Effect sizes of genome-wide significant loci on appetite traits before and after adjustment for BMI at age 8. Filled circles represent crude effect sizes (β) and open diamonds represent BMI-adjusted effect sizes, with horizontal lines indicating 95% confidence intervals derived from standard errors. Loci are grouped by appetite domain: food responsiveness (FR, orange), satiety responsiveness (SR, blue), and food fussiness (FF, green). The percentage attenuation in effect size following BMI adjustment is shown on the right. TMEM18 (cond.) denotes the conditional signal at the TMEM18 locus, independent of the primary signal.
